# Risk assessment via layered mobile contact tracing for epidemiological intervention

**DOI:** 10.1101/2020.04.26.20080648

**Authors:** Vishwesha Guttal, Sandeep Krishna, Rahul Siddharthan

## Abstract

There is strong interest globally amidst the current COVID-19 pandemic in tracing contacts of infectious patients using mobile technologies, both as a warning system to individuals and as a targeted intervention strategy for governments. Several governments, including India, have introduced mobile apps for this purpose, which give a warning when the individual’s phone establishes bluetooth contact with the phone of an infected person. We present a methodology to probabilistically evaluate risk of infection given the network of contacts that individuals are likely to encounter in real life. Instead of binary “infected” or “uninfected” statuses, an infection risk probability is maintained which can be efficiently calculated based on probabilities of recent contacts, and updated when a recent contact is diagnosed with a disease. We demonstrate on realistic networks that this method sharply outperforms a naive immediate-contact method even in an ideal circumstance that all infected persons are known to the naive method. We demonstrate robustness to missing contact information (such as when phones fail to make bluetooth contact or the app is not installed). We show, within our model, a strong flattening of the infectious peak when even a small fraction of cases are identified, tested and isolated. In the real world, where most known-infected persons are isolated or quarantined and where many individuals may not carry their mobiles in public, we believe the improvement offered by our method warrants consideration. Importantly, in view of widespread concerns on privacy and contact-tracing, our method relies mainly on direct contact data that can be stored locally on users’ phones, and uses limited communication via intermediary servers only upon testing, mitigating privacy concerns.

## Introduction

The COVID-19 coronavirus pandemic, which has expanded from China in December 2019 to affect almost every country in the world by now (April 2020), has led to a strong interest in non-pharmacological interventions to curtail spread. Early efforts in China, Singapore, South Korea and other countries involved extensive testing as well as identification and isolation of contacts of infected individuals and mobile-based alerts [13]. Several governments have also experimented with mobile contact-tracing applications. At a basic level, these applications enable a mobile phone to communicate with other mobile phones via bluetooth, and warn the owner when contact has been made with an infected person. An example is India’s Aarogya Setu (“health bridge”) app^1^. Previously, such apps were developed in China, Singapore, and South Korea [10], and are under development in countries including France [7], the USA [10], and the UK [12] and elsewhere. Privacy concerns have been raised globally, and privacy-sensitive protocols have been proposed [4]. Meanwhile Google and Apple have announced a partnership to develop contact-tracing infrastructure for inclusion in their basic mobile software stacks, promising to respect privacy and security. [8]

Unfortunately most current apps are closed-source with opaque mechanisms, but as far as is documented by them, they rely on direct contacts with known infected individuals, and possibly on past direct contacts with individuals who subsequently became diagnosed as infected.

Here we present a probabilistic framework to assess an infection risk based on risk factors of contacts, whether positive or not. In this scheme, every individual has a risk factor based on their contact history. We demonstrate via simulations that this method strongly outperforms a naive method based only on direct contacts. Given some epidemiological assumptions and approximations, our calculation is rigorous but can be performed locally on a mobile phone using only the owner’s risk factor and the risk factor of the contact. Contact history, too, can be stored on the mobile phone and need not be shared with a server. Only one crucial step, of updating risk factors of recent contacts based on subsequent diagnoses, may require the use of a server. This too may probably be made secure, but we do not discuss security issues here, only efficiency of identifying likely infected individuals (who could, for example, be asymptomatic but spreaders).

## Methods

### Risk factor evaluation

We make the approximation that the person-to-person transmission probability is a constant per contact, *p_t_*. This parameter is related to the infection rate in the SEIR compartmental model, discussed in a later subsection.

If individual A, who is uninfected, makes one contact with individual B, who is infected, then A gets a probability *p_t_* of being infected after that contact. But we can also consider the case when B has a probability *p_B_* of being infected; then the probability of A being infected after the contact is *p_B_p_t_*.

The most general scenario is that neither A nor B were uninfected, but had previous probabilities *p_A_* and *p_B_* of being infected. Then A is now infected with a new probability 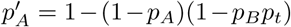. Note that the bracketed terms are the probabilities of being originally uninfected, and of remaining uninfected after the contact. Similarly, we update B’s probability too, to 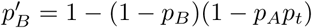. Note that if *p_B_* = 0 then *p_A_* is unchanged, while *p_B_* updates to *p_A_p_t_*, as expected; and vice versa.

We therefore propose the following algorithm to update individual probabilities, or risk factors, of being infected.

1. Each individual has a unique ID (for example, mobile phone number).
2. Each individual’s infection probability is initialized in some way based on prior knowledge and self-reporting. This could be done via a questionnaire upon installing the app. The vast majority will be initialized to zero.
3. Each individual’s app maintains a list of ALL contacts in the past *m* days (*m* ≈ 14, estimated upper bound of the incubation time; contacts from earlier are unlikely to affect current infection status). With each contact is included a list of all meeting times with that contact. This is required for a thorough update of probabilities, as discussed below.
4. Every time two individuals A and B are in proximity, their mobile apps exchange their infection statuses *p_A_* and *p_B_*. The update is made as above: 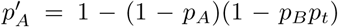 and inline equ 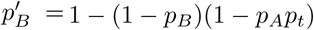
5. **Test update propagation:** If person A on B’s contact list tests positive, then *p_A_* is updated from its previous value to 1. But since A was probably already infected during their contact (given the long incubation time of the virus), *p_B_* needs to be updated too, to 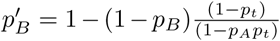 (where *p_A_* in the denominator is the previous value for A, and the new value for A is 1). But now we need to update every C whom B met subsequent to meeting A (that is, B’s *last* meeting with C is more recent than B’s *first* meeting with A). The formula for this is 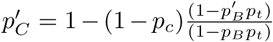. And, again, every contact of C who was met subsequent to C’s first meeting with B, and so on. In our simulation we accomplish this via a recursive function, avoiding cycling back to a previous contact by passing an “ignore list” of contacts in each function call. Additionally, if a contact was met multiple times in the relevant timeframe, an update is performed the same number of times, since each meeting carried a risk.
6. Each individual A’s contacts older than *m* days drop off the contact list, but this does not change *p_A_*. That is, if B, who last met A more than *m* days ago, is diagnosed infected, this is unlikely to require updating *p_A_*.
7. Individuals who are recovered are marked as as immune. They play no further part in our simulation. In the real world, some instances of re-infection within a short time frame have been noted ([5], and additional cases reported in news media).

### Simulation

We simulate an agent-based model on a network, in which agents interact stochastically over time and are categorized as ‘‘susceptible”, ‘‘exposed”, ‘‘infectious”, and ‘‘recovered”. These are the categorizations of the compartmental SEIR model in epidemiology, discussed and compared in the next subsection. We implement the risk update algorithm on the same agent-based framework and compare the risk profile predicted by our algorithm with the actual infections of the agent-based simulations.

We initialize a population of size N, whose individuals are nodes on a weighted network. The network represents all *possible* contacts in this population: a link indicates two people who may make a contact, and the weight of the link is the probability of their making a contact at a given time. We consider random networks with uniform degree distribution and uniform link weights, Barabàsi-Albert-structured networks, and networks with family structures and small-world features. Our results are consistent across all these structures. A key parameter of the network, used below, is the average number of contacts per node, defined as

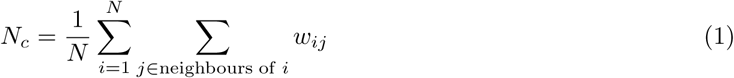

where *w_ij_* is the weight of the link between *i* and *j*.

Individuals are marked as susceptible (S), exposed (E—infected but not yet infectious), infectious (I) and recovered (R, assumed immune to future infection). The simulation is initialized with all individuals being uninfected (susceptible) except a small number (eg, 10 out of 10,000) who are infectious. With each individual is associated a probability, which is initialized to 1 for infectious individuals and 0 for others.

In each pass of the simulation, which we call an “epoch”, every link on the graph is sampled once, and a contact is made with probability equal to the weight of the link. So links weighted 1 (such as family links) are always sampled, while other links may be rarely sampled. After each contact between a pair of individuals, if one is infectious and the other is susceptible, the other is marked “exposed” with a probability *p_t_*. For contacts other than S-I, nothing is done.

At the end of each epoch, each individual is sampled for their status. Exposed individuals become infectious with a probability p_e_ and infectious individuals recover with a probability *p_r_*.

*In parallel with this*, at each contact, the probability scores of individuals making contact are updated as described above in “Risk factor evaluation”. This is done for each sampled pair of contacts if at least one has a non-zero probability score.

An example of a possible simulation is in figure 1.

**Figure 1:**
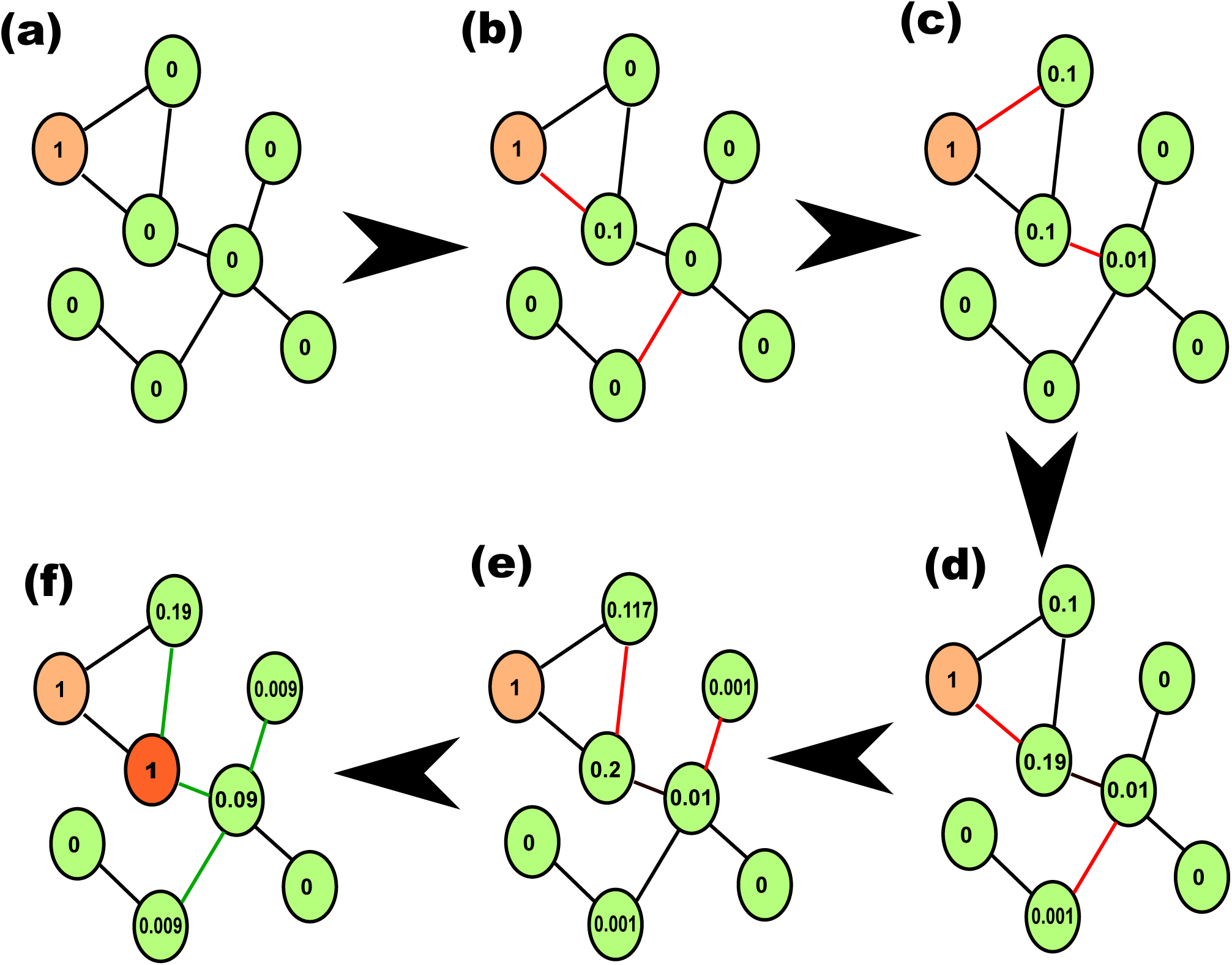
An example of a possible simulation, with *p_t_* = 0.1. Numbers in ovals are probability values. The ”infected” status values are not shown here but are updated stochastically in parallel with the probabilities. (a) Network with one individual initially infected. (b)-(e) Red links indicate contacts, and probabilities of respective nodes are updated. (f) A second individual is diagnosed infected (dark red) and that individual’s probability value is changed to 1.0, and the chain of previous contacts is updated (green links).

We also keep track of a “naive probability” for each individual, which consists simply of updating

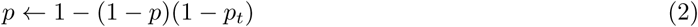

every time a known susceptible individual meets a known infected individual. We call this the “naive oracle” approach, since this algorithm does not consider contact with people who have a risk factor, only with truly infectious people; but knows the true infectious status of the contacted person. In the real world, this is known for only a fraction of infectious people.

Thus, the parameters of the simulation are *p_t_*, *p_e_* and *p_r_*. However, these are in turn derived from other parameters as follows: *R_0_* is the “basic reproduction number” (see next subsection); *M_d_* is the number of epochs that constitute one day; *d_e_* is the average exposure time in days (the average number of days to turn infectious); *d_r_* is the average recovery time in days. From these, we get 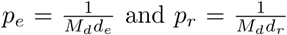. Finally, *p_t_* is determined from *R_0_* and *N_c_* as in the next section (equation 9).

### SEIR epidemiological model

The SEIR compartmental model in epidemiology, [16], without vital dynamics (i.e., without births and deaths in the population), is usually written as

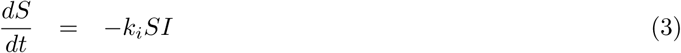

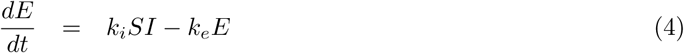

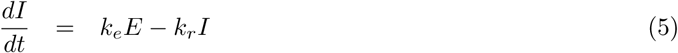

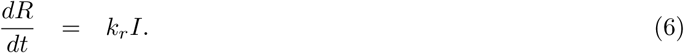

Here *S* is the number of susceptible individuals in the population, *E* is the number of individuals exposed to infection but not yet infectious, *I* is the number of infectious individuals, *R* is the number of recovered individuals. The rate of infection (per person) is *k_i_*, exposed individuals become infectious at rate *k_e_*, and the recovery rate is *k_r_*. These equations conserve the total population *S* + *E* + *I* + *R*. Within this model recovered individuals are permanently recovered (though deaths are not included here, individuals who die of the disease may also be counted in R since they are no longer infectious).

We seek to estimate parameters of our simulation from epidemiological measurements in the real world. A key epidemiological parameter is *R_0_*, the “basic reproduction number” or the average number of individuals infected by any individual while infectious. It can be shown easily[16] that

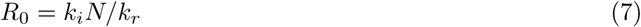

where *N* is the total population. This is valid for the SIR model as well as the SEIR model without births and deaths. This assumes a “well-mixed” system, but otherwise the same equation is commonly used with *N* being an “effective population”.

In terms of individual contacts and contact rates, we can alternatively write

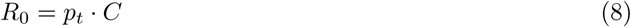

where *p_t_* = transmission probability per contact as above, and *C* is the total number of contacts while the patient is infectious. *C* is equal to the rate of contact *R_c_* (per epoch, say) times the average recovery time (also in epochs). So if the contact rate is 100 contacts per epoch, and the recovery time is 10 epochs, then *C* = 1000, and if *R*_0_ = 2, then *p_t_* = 0.002.

The average recovery time is 1/*k_r_*, so comparing the two definitions of *R*_0_ (equations 7 and 8), we can identify *k_i_N* = rate of infection = rate of contacts × probability of infection per contact = *R_c_p_t_*. Therefore, *p_t_* = *R*_0_*k_r_*/*R_c_*. If, for example, *R*_0_ is empirically estimated as 2, the recovery time is taken to be 10 days, and the average rate of contact per day is 100, then we estimate *p_t_* as 0.002. More generally, we take the rate of contact per *epoch* to be exactly equal to the average number of contacts per link, *N_c_* (equation 1). Then we have

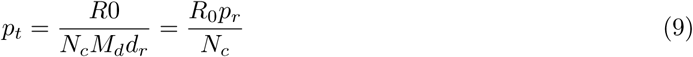

which we use in the network simulation.

### Gillespie simulations

To validate the accuracy of our sampling simulation, we also simulate the spread of the infection using the Gillespie algorithm. At any given time the state of the system is fully specified by the state of each individual; for the *i*th individual, its state *s_i_* ϵ {S (susceptible), E (exposed), I (infectious), R (recovered)}. Let *N_S_*(*t*),*N_E_*(*t*),*N_I_*(*t*),*N_R_*(*t*) be the number of individuals in each of these states at time *t*. Time is again divided into epochs. At the beginning of each epoch the network links are sampled as above. Let *C_ij_* (*t*) = 1 denote that there is a contact between individuals *i* and *j* in the epoch within which time *t* falls; *C_ij_* (*t*) = 0 if there is no contact between these two in that epoch. Then the Gillespie algorithm is run until the time of the next epoch as follows:

1. Assign the start time of the epoch to the variable *t*
2. List all possible “events” that can occur which will change the state of the system: there will be 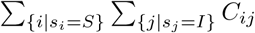 infection events, *N_E_* exposed-becoming-infectious events, and *N_I_* recovery events possible, for a total of *n* events. For each of these, compute the rate or probability per unit time for that event to occur, denoted *a_j_* for event *j*, *j* ϵ {1, 2,…, *n*}.
3. Let 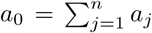. Let Δ*t* be a random number sampled from the exponential distribution with mean 1/*a*_0_.
4. If *t* + Δ*t* is larger than the time of the next epoch, set *t* equal to the time of the next epoch and end the Gillespie run, resample the network and run the Gillespie algorithm for the next epoch.
5. Otherwise, update *t* to be equal to *t* + Δ*t*, and choose one event to occur at that time (event *j* is chosen with probability *a_j_*/*a*_0_). Update the state of each individual if the event has changed it, along with *N_s_*(*t*), *N_e_*(*t*), *N*/(*t*), *N_R_*(*t*) (for example, if individual *i* infects individual *j*, then *S_j_* (*t*) = *E*, *N_E_* is incremented and *N_S_* is decremented by one).
6. Go to step 2.

This process is repeated for as many epochs as desired, usually until the number of exposed and infected individuals has fallen to zero. This gives the distributions *N_S_*(*t*), *N_E_*(*t*), *N_I_*(*t*) and *N_R_*(*t*) as functions of time, for comparison with the network sampling simulation.

## Results

### Complete network: Comparison with well-mixed SEIR model

Figure 2 (a) shows a simulation on 2000 nodes, each node connected to all others with weight 1.0. For this “fully connected” or “complete” network, the SEIR model, with parameters determined as in the figure caption, shows excellent agreement with our simulation. The unrealistic assumption here is that each member of the population meets each other member exactly once per epoch; we exhibit it to demonstrate the agreement with a well-mixed SEIR model, but do not explore this network further.

**Figure 2:**
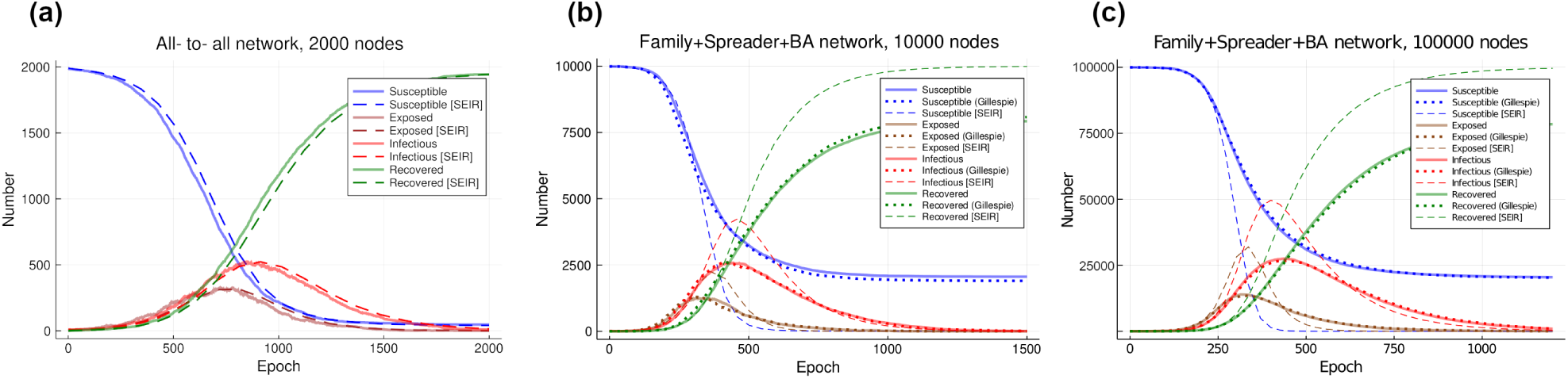
(a) A comparison of our network simulation on 2000 nodes, with every node connected to every other node with weight 1.0, with the compartmental SEIR model. The parameters taken for the network simulation were: *R*_0_ = 4.0, *M_d_* = 10, *p_e_* = 1/10, *p_r_* = 1/20, *p_t_* ≈ 1.0 × 10^−5^ (from equation 9). For the SEIR integration, since all rates are small, we use *k_e_* = *p_e_*/*M*_d_ = 0.01, *k_r_* = *p_r_*/*M_d_* = 0.005, and *k_i_* = *p_i_*. The simulation over 2000 epochs (200 days) agrees very well with the SEIR solution. (b) Simulation on a 10000-node network that includes family units (size 1–5; link weight 1), spreaders (1% of total, 1000 links each, link weight 0.1 each) and links added via the Barabási-Albert method (link each node to a random new node with probability proportional to current coordination number of new node; link weight 0.1). This network has 183,599 links. We used *M_d_* = 10 epochs. The average contact of each node is 3.77/epoch, ie 37/day. Parameters were: *R_0_* = 3, *d_e_* = 5 days (5 epochs), *d_r_* = 15 days(150 epochs), *p_t_* = 0.0053 (calculated from above). The simulation over 1500 epochs (150 days) agrees well with an independently implemented Gillespie simulation, but disagrees with the SEIR prediction. Shown is SEIR for effective population size *N_c_* = 4000, which gives the best fit but is hard to justify. (c) Simulation on a 100,000 node network with 1,948,709 links, layered similarly to the network in (b), except that the BA links have weight 0.05, with the same parameters except *p_t_* = 0.0069 (calculated). The SEIR plotted is for effective population 23,000.

### Realistic network: Comparison with SEIR model and Gillespie simulation

We construct a more realistic network as follows. We initialize a random network with family links, such that every individual belongs to a family of size 1, 2, 3, 4, or 5 (with relative probabilities 0.2, 0.3, 0.3, 0.1, 0.1). Family networks are complete, i.e., all members are connected to all others with weight 1. Then, 1% of the individuals are randomly selected, and each is connected to 1000 other individuals, with weight 0.1. These are meant to indicate “spreaders”, that is, people such as shop attendants and receptionists who have a large number of daily contacts that vary every day. Further links are added via a modification of the Barabási-Albert (BA) algorithm[2], that is, each node is attached to a randomly selected other node with weight 0.1. In the BA algorithm the new node is selected from all other nodes with a probability proportional to their coordination numbers. For efficiency, we first select 1000 random other nodes, and then select from those using the BA method.

Figure 2 (b) shows the result of simulation on this network over 1500 epochs (150 days). The SEIR simulation shown is with an effective population size of 4,000, which has no justification but offers a better fit compared to either the total population (10,000) or the average contact number (37). It is known that the rate of spread of an infection on a network depends on its structure and not just on the average contact number [17], so it is not surprising that the best choice of effective population is different from the average contact number. In comparison, a Gillespie simulation on the same network agrees very well.

Figure 2 (c) shows a simulation on 100,000 nodes, with parameters as in the caption. The SEIR here is plotted for an effective population of 23,000, and again fits poorly while the Gillespie agrees well.

### Growth of probabilities

While the object of this exercise is not to predict overall numbers in the population, but to identify individuals most at risk, it is of interest to see how this probability varies with the growth in infectious individuals. Figure 3 (left) plots the “total probability” (the sum over all individuals of individual probabilities, which would crudely be the expected number of infected individuals), as well as the number of individuals with *p* exceeding 0.5, 0.6, 0.7, 0.8, 0.9. The total grows much faster than the epidemic, as do each of the fractional curves, but the latter start their growths at different times that roughly track the growth of infectious cases.

**Figure 3:**
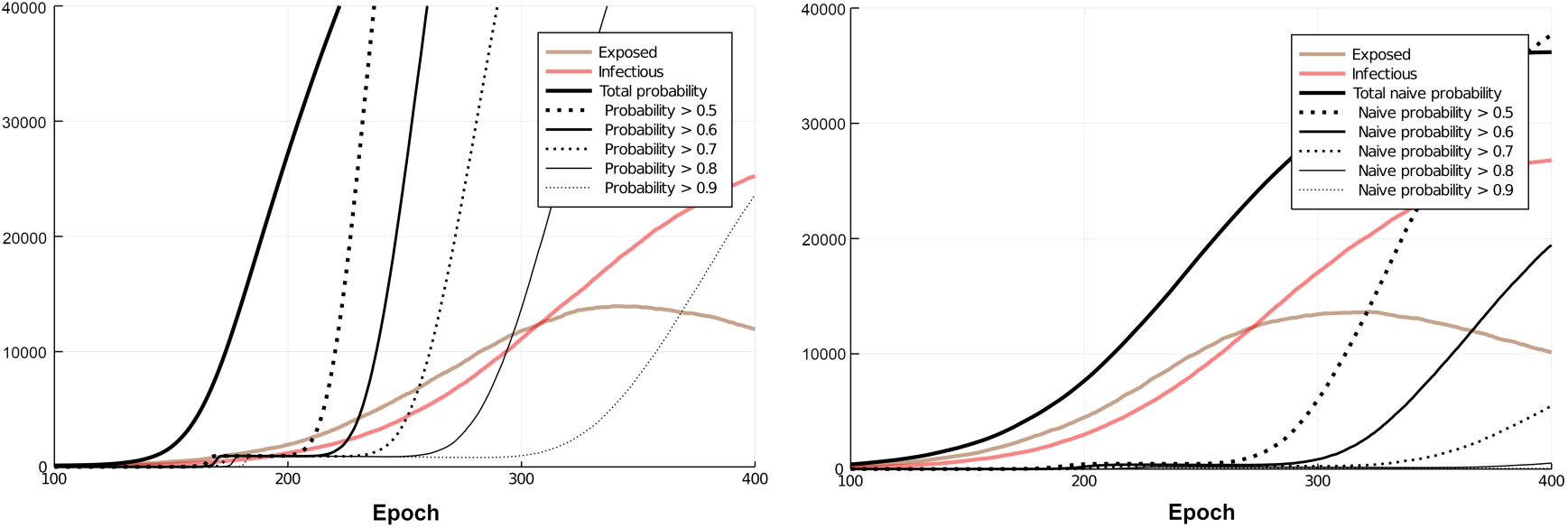
(left) The “total probability” (*p* summed over all 100,000 individuals) as well as the fraction of individuals over 0.5, 0.6, 0.7, 0.8, 0.9 in probability. (right) Similar plot for the naive probability.

Figure 3 (right) does the same for the naive probability. Here the total probability tracks the infectious total more closely, but the fractional curves appear to increase at roughly the same epoch. This perhaps gives some intuition for the performance of our method compared to the naive method as measured by positive rate vs false positive rate, or precision vs recall, discussed below.

Notably, at this time we have no “recovery” for probabilities, other than the testing-and-resetting mechanism discussed further below, not used in this figure. The methodology is expected to be most useful in early stages of an epidemic.

### Effectiveness of probabilistic prediction compared to naive methods

On the network exhibited in Figure 2 (c), we compare the predictions of our probabilistic method and the “naive oracle” method at epochs 200, 250 and 300 (representing 1815, 6479 and 14126 infections respectively out of 100,000). Subfigures 4 (a), (b), (c) show receiver operating characteristic (ROC) curves, and subfigures (d), (e), (f) show precision-recall curves (PRC). Also shown is the result for a random assignment of probability in (0, 1) for each individual. In all cases probabilistic method clearly outperforms the naive oracle, even though the oracle has the strong advantage of knowing whether an individual in an encounter is truly infected or not. These are plotted by calculating true positive rates (also called recall), false positive rates, and precision varying the threshold *p* used to predict an individual’s infective state. A true positive is an infected patient who is predicted to be infected (*p* above threshold); a false positive is an uninfected patient predicted infected; a true negative is an uninfected patient predicted uninfected; a false negative is an infected patient predicted uninfected. If the total numbers of these are respectively *TP*, *TN*, *FP* and *FN* then

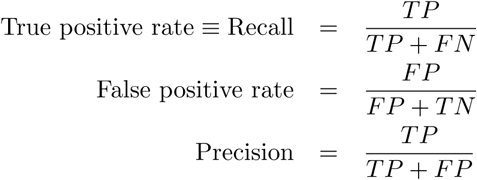

**Figure 4:**
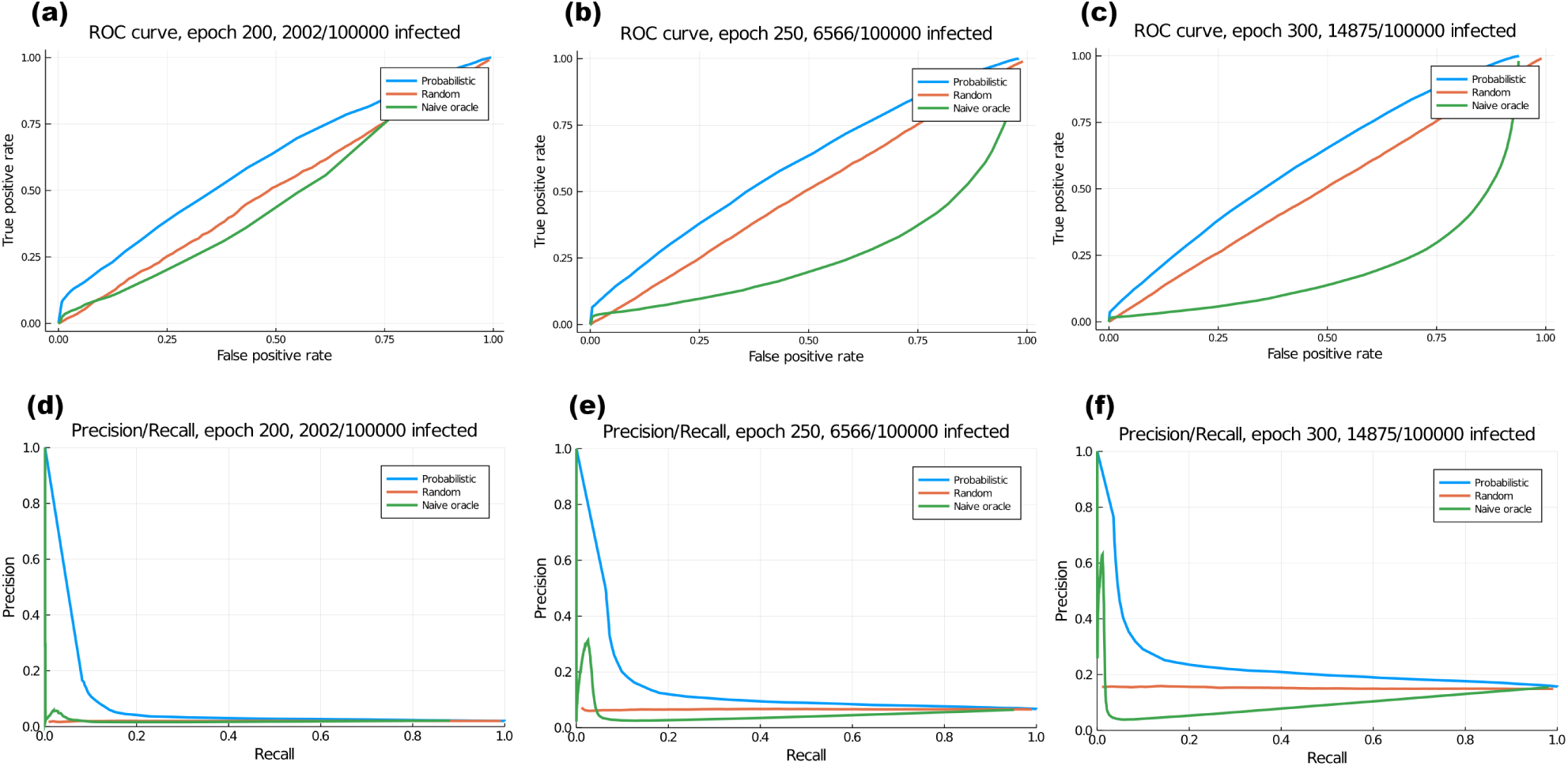
(a), (b), (c): ROC curves for our probabilistic prediction of infections, a random allocation of probabilities, and the naive oracle method, at epochs 200, 250 and 300 on the network in figure 2 (c). (d), (e), (f): Precision recall curves at the same epochs.

So, at all epochs shown, the probabilistic method has a TPR of about 67% at a FPR of 50%; the naive oracle performs at less than 50% TPR at 50% FPR in all cases, and its performance grows worse at later epochs (as the infection spreads.) Since suspected individuals will be tested via accurate RT-PCR tests, we feel this FPR rate is acceptable, especially given the effectiveness of a testing+isolation strategy that tests even a small fraction of risky individuals (next section)

### Effectiveness of testing and isolation of patients

The naive oracle above is assumed to know the status of every covid-19 positive patient. Also, we update naive probabilities only on contact between infectious and uninfected patients.

For a more realistic comparison with the real world operation of these methods, we can simulate “testing” of patients, after which they are marked “tested positive” (known infectious) or negative (susceptible).

We implement testing at each epoch by selecting a predetermined fraction of all individuals with a high probability to be tested; the test simply looks at their true infected status. If truly infected, they are marked “tested positive”, their probabilities and naive probabilities are set to 1, and non-naive probability updates are propagated to their contacts (as in figure 1 (f)). If they test negative, they are marked susceptible, and their probabilities are set to zero and their contact updates propagated.

We modify the naive probability tracing to only consider contacts with known-infectious (tested) cases, and to update as in equation (2) for each such contact (regardless of the status of the contacter.) The sophisticated tracing goes on as before, but is aware of known-infectious contacts because their probabilities are set to 1 (but does not deal with them in any special way).

The results are shown in figure 5, for a test threshold of *p* = 0.9 and rates of 5% and 1% of above-threshold cases tested. Interestingly, it appears that this non-oracular naive method is only able to achieve a very small recall (TPR) regardless of precision. Also, overall performance of the probabilistic method is reduced in the presence of testing, perhaps because probabilities after negative tests are being set to zero even though they could be in risky populations and liable to be infected.

**Figure 5:**
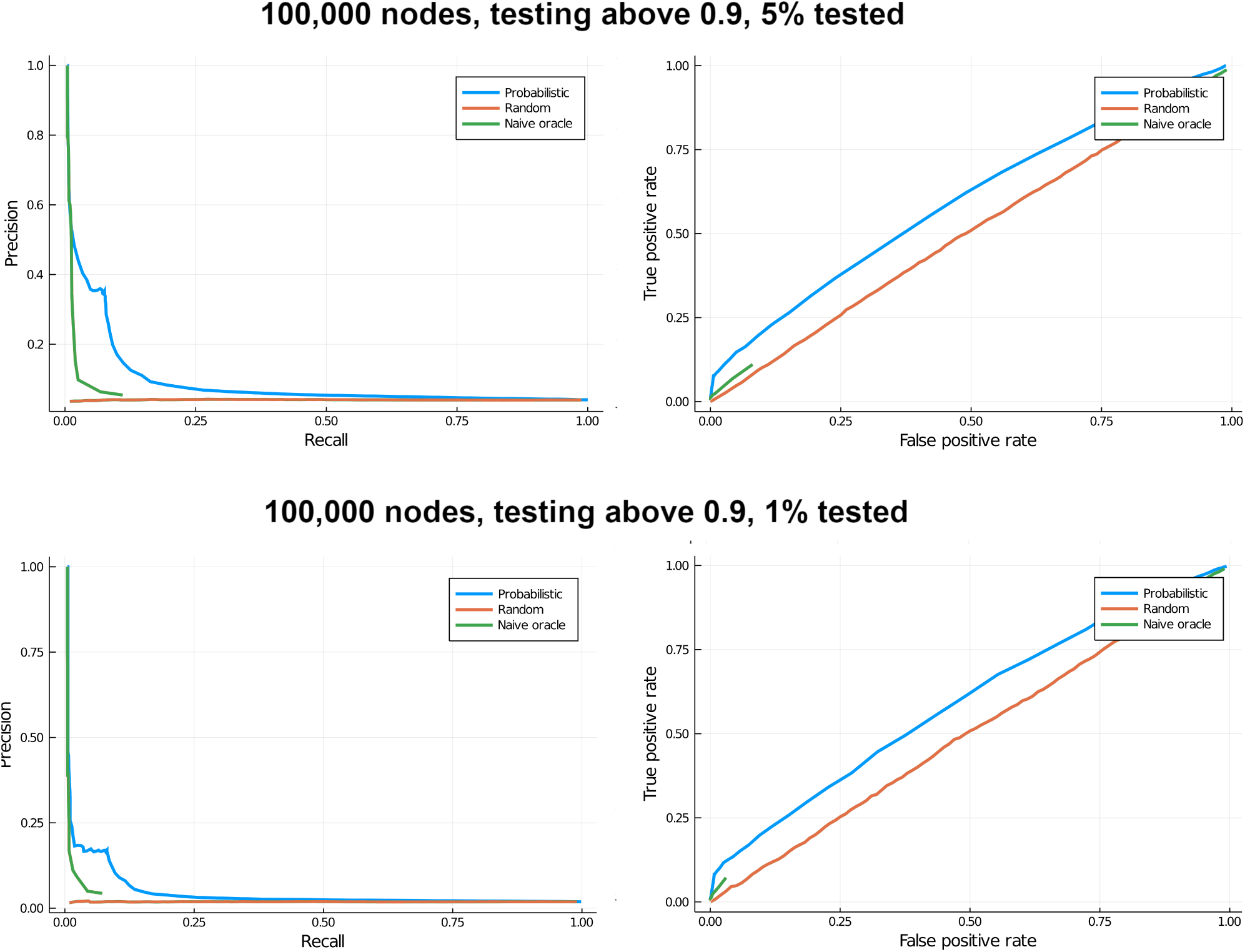
ROC and Precision-Recall curves with testing for a non-oracle naive method, which is aware only of tested-positive individuals, and the probabilistic method.

We can also isolate tested-positive patients, by weakening their links to all their contacts. Figure 6 (a) shows the effect of this, for a testing threshold of 0.8 (individuals whose probability exceeds 0.8 are tested), for testing percentages of 10% and 20% of risky individuals, and with a weakening of links to 10% of their previous value. (For a population of 100,000, at epoch 300 in our simulation, about 2,200 individuals have *p* > 0.8, so 10% testing here means testing 220/100,000 individuals, or 0.2% of the full population.) This suggests that a test rate of even 10% has a very strong effect in flattening the curve. However, though this suggests the effectiveness of testing and isolation (which has been widely noted [1, 14, 3] and is being practised by most countries), we caution against drawing quantitative conclusions from our model.

**Figure 6:**
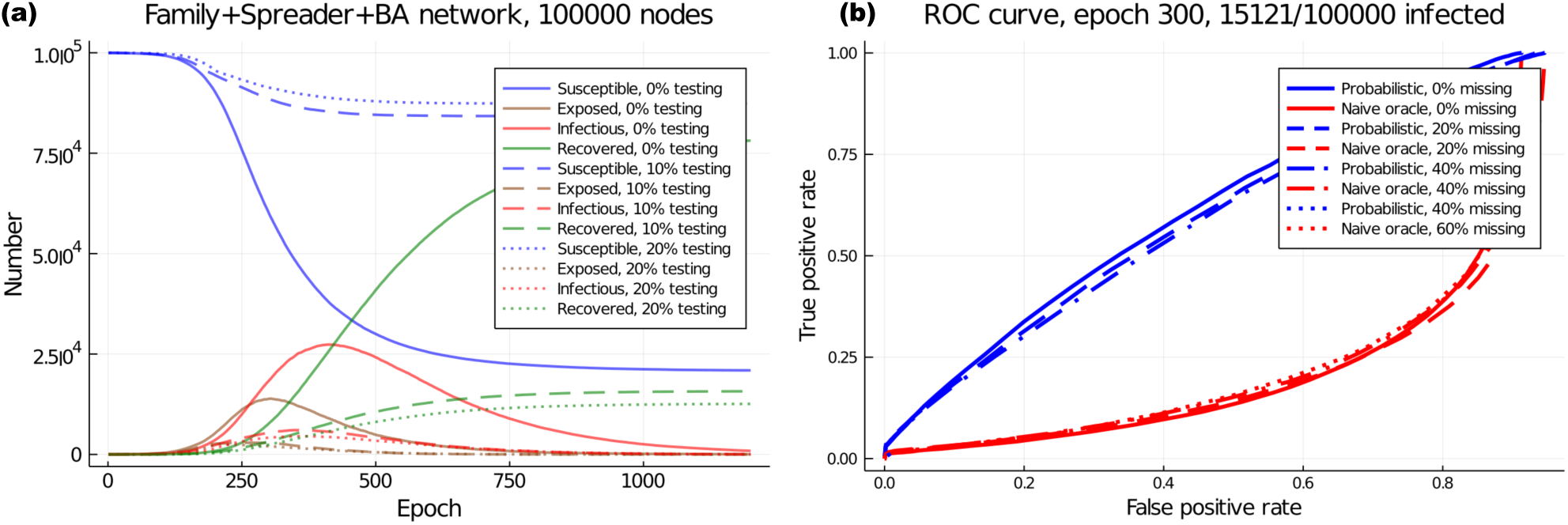
Effect of testing, and of missing contacts

### Lossy data

With mobile tracking, it is likely that several individuals will not be carrying their mobile or will not have the app installed, therefore the probabilistic updates will not occur. Figure 6 (b) shows the effect of such missing contacts, implemented by randomly ignoring updates with a given probability. This appears to have negligible effect for up to 60% loss in contacting (40% successfully recorded contacts).

## Discussion

Several authors have discussed the possibility of tracing contacts of recently infected patients via mobile phones, with the goal of isolating them. This is argued [9, 3] to be an effective way to control the outbreak and build “digital herd immunity”. We demonstrate in an agent-based simulation on a network that our method is a better predictor, based on TPR/FPR or precision/recall, of truly infected patients compared to a naive first-contact-based prediction, even in an ideal case where the naive method is an “oracle” that always knows the true status of the contact. Our results are robust to loss in detection of contacts, which is expected to be significant in real life. Our simulations show that testing only the most probable individuals (*p* > 0.8) and isolating them (reducing link strength by a factor of 10) strongly flattens the curve of infection. Though we have tried to make our network structure realistic, the real world has several complications over a simulation; nevertheless we expect these results to hold qualitatively.

Several authors have also raised privacy concerns [11, 6] over mobile contact tracing, and proposed privacy protocols to handle this [4]. We do not directly address privacy concerns here. However, our method requires most contact information to be stored only on the users’ own mobile phone. This information only consists of the contact’s mobile number, number of meetings, and times of first meeting and last meeting. Infection probability information is exchanged via bluetooth at the time of contact, but is used only to update one’s own probability and need not be stored. Only one step, the “update contacts” procedure that propagates the change in diagnosis of an individual to the individual’s contacts, and the contacts’ contacts, recursively, requires the means for one mobile phone to communicate to another post-contact. This likely requires the use of an intermediary server, but this use is limited and privacy concerns can be mitigated by using an encrypted protocol and deleting communication request data once the request is carried out.

Overall, our probabilistic contact tracing framework appears to outperform the naive method significantly, whether implemented as an “oracle” that knows all truly infected individuals, or implemented with a testing framework to recognize only positively-tested individuals. While it can be used to identify immediate contacts of a tested individual, it can go further to identify at-risk individuals in the wider population, while also substantially taking care of privacy concerns.

While we focus on the SEIR disease model, more complex models featuring asymptotic individuals, different levels of symptomatic individuals, limited recovery period (recovered individuals becoming susceptible after a time), etc, can be considered and are being considered for COVID-19 [15] ^2^. Our simulation framework can be modified easily for such cases too, as well as for bigger and more geographically structured networks. We plan to explore such complexities in future work.

## Data Availability

The manuscript describes an algorithm for contact tracing and a simulation. Code for this is available at https://github.com/rsidd120/EpiTracSim/

## Acknowledgements

The authors would like to acknowledge early discussions with Farhat Habib and Mukund Thattai, and useful feedback from Gautam Menon and Leelavati Narlikar, as well as general and ongoing discussions with the Indian Scientists’ Response to COVID-19 group^3^. VG acknowledges support from DBT-IISc partnership program. SK is funded by the Simons Foundation, and the Department of Atomic Energy, Government of India, under project no. 12-RD-TFR-5.04-0800. RS is funded by the Computational Biology project at his institute, from the Department of Atomic Energy, Government of India.

## Availability

The network generation and simulation code is available at https://github.com/rsidd120/EpiTracSim.

## Footnotes

1 https://www.mygov.in/aarogya-setu-app/

2 Ongoing study at https://indscicov.in/indscisim

3 https://www.indscicov.in/

## References

[1] Roy M Anderson, Hans Heesterbeek, Don Klinkenberg, and T Déirdre Hollingsworth. How will country-based mitigation measures influence the course of the COVID-19 epidemic? The Lancet, 395(10228):931–934, 2020.

[2] Albert-László Barabási and Réka Albert. Emergence of scaling in random networks. science, 286(5439):509–512, 1999.

[3] Vir B Bulchandani, Saumya Shivam, Sanjay Moudgalya, and SL Sondhi. Digital Herd Immunity and COVID-19. *arXiv preprint arXiv:2004–07237*, 2020.

[4] Justin Chan, Shyam Gollakota, Eric Horvitz, Joseph Jaeger, Sham Kakade, Tadayoshi Kohno, John Langford, Jonathan Larson, Sudheesh Singanamalla, Jacob Sunshine, et al. PACT: Privacy Sensitive Protocols and Mechanisms for Mobile Contact Tracing. *arXiv preprint arXiv:2004.03544*, 2020.

[5] Dabiao Chen, Wenxiong Xu, Ziying Lei, Zhanlian Huang, Jing Liu, Zhiliang Gao, and Liang Peng. Recurrence of positive SARS-CoV-2 RNA in COVID-19: A case report. International Journal of Infectious Diseases, 2020.

[6] Hyunghoon Cho, Daphne Ippolito, and Yun William Yu. Contact tracing mobile apps for covid-19: Privacy considerations and related trade-offs. *arXiv preprint arXiv:2003.11511*, 2020.

[7] Romain Dillet. France is officially working on ‘Stop Covid’ contact-tracing app. TechCrunch, Apr 2020.

[8] Adam Clark Estes and Shirin Ghaffary. Apple and Google want to turn your phone into a Covid-tracking machine. Vox, Apr 2020.

[9] Luca Ferretti, Chris Wymant, Michelle Kendall, Lele Zhao, Anel Nurtay, Lucie Abeler-Dörner, Michael Parker, David Bonsall, and Christophe Fraser. Quantifying SARS-CoV-2 transmission suggests epidemic control with digital contact tracing. Science, 2020.

[10] Shirin Ghaffary. How China, Singapore, Taiwan, and other countries have been using technology to battle Covid-19. Vox, Apr 2020.

[11] Marcello Ienca and Effy Vayena. On the responsible use of digital data to tackle the COVID-19 pandemic. Nature Medicine, pages 1–2, 2020.

[12] Leo Kelion. Coronavirus: NHS contact tracing app to target 80% of smartphone users. BBC News, Apr 2020.

[13] Justin McCurry, Rebecca Ratcliffe, and Helen Davidson. Mass testing, alerts and big fines: the strategies used in Asia to slow coronavirus. The Guardian, Mar 2020.

[14] Wolfgang Preiser, Gert Van Zyl, and Angela Dramowski. COVID-19: Getting ahead of the epidemic curve by early implementation of social distancing. SAMJ: South African Medical Journal, 110(4):0–0, 2020.

[15] Mihir Arjunwadkar Dhiraj Kumar Hazra Pinaki Chaudhuri Sitabhra Sinha Gautam I Menon Anupama Sharma Vishwesha Guttal Snehal Shekatkar, Bhalchandra Pujari. INDSCI-SIM A state-level epidemiological model for India, 2020. Ongoing Study at https://indscicov.in/indscisim.

[16] Pauline van den Driessche. Reproduction numbers of infectious disease models. Infectious Disease Modelling, 2(3):288–303, 2017.

[17] Duncan J Watts and Steven H Strogatz. Collective dynamics of ‘small-world’networks. nature, 393(6684):440, 1998.

